# In-hospital Outcomes of Acute Ischemic Stroke in Atrial Fibrillation Patients on Anticoagulation: A Retrospective Analysis

**DOI:** 10.1101/2023.12.21.23300418

**Authors:** Moises A Vasquez, Litsa K. Lambrakos, Alex Velasquez, Jeffrey J. Goldberger, Raul D. Mitrani

**Author notes:** Corresponding author: Address: 1611 NW 12th Ave, C-600D, Miami, FL 33136 Department of Internal Medicine, University of Miami Miller School of Medicine/Jackson Memorial Hospital, Miami, Florida, USA. Authors declare no conflict of interest.

## Abstract

**Background:** Long-term anticoagulation (AC) therapy reduces the risk of stroke in patients with Atrial Fibrillation (AF). Data on the impact of AC on in-hospital stroke outcomes is lacking.

**Methods:** The National Inpatient Sample was used to identify adult inpatients with AF and a primary diagnosis of ischemic stroke between 2016 and 2020. Data was stratified between patients on AC users and nonusers. A multivariate regression model was used to describe the in-hospital outcomes, adjusting for significant comorbidities.

**Results:** A total of 655,540 patients with AF and a diagnosis of ischemic stroke were included, of which 194,560 (29.7%) were on long-term AC. Patients on AC tended to be younger (mean age, 77 vs 78), had a higher average CHA2DS2VASc score (4.48 vs 4.20), higher rates of hypertension (91% vs 88%), hyperlipidemia (64% vs 59%), and heart failure (34% vs 30%) compared to patients not on long-term AC. Use of AC was associated with decreased in-hospital mortality (aOR [95% CI]: 0.62 [0.60 - 0.63]), decreased stroke severity (mean NIHSS, 8 vs 10), decreased use of tPA (aOR 0.42 [0.41 - 0.43]), mechanical thrombectomy (aOR 0.85 [0.83 - 0.87]), hemorrhagic conversion (aOR 0.69 [0.67 - 0.70]), gastrointestinal bleeding (aOR 0.74 [0.70 - 0.77]), and discharge to skilled nursing facilities (aOR 0.90 [0.89 - 0.91]), compared to patients not on AC (P<0.001 for all comparisons).

**Conclusion:** Among patients with AF admitted with acute ischemic stroke, AC use prior to stroke was associated with decreased in-hospital mortality, decreased stroke severity, decreased discharge to SNF, and fewer stroke-related and bleeding complications.

## Introduction

Atrial Fibrillation (AF) is associated with an increased risk of ischemic stroke, systemic embolism, and death (1). In the USA, about 10%–12% (∼80000) of ischemic strokes occur in the setting of AF and carry higher morbidity and mortality than those occurring in patients without AF (2). Oral anticoagulation (AC) with direct oral anticoagulants (DOACs) or vitamin K antagonists (VKA) have shown to substantially reduce the risk of stroke. Nevertheless, despite these advancements in stroke prevention, strokes in patients with AF continue to be a significant burden. Recent studies show that 22% to 36% of ischemic strokes in patients with AF occur while on anticoagulation(3,4). Furthermore, patients with AF that have had a prior stroke are at a higher risk of stroke recurrence, even while on AC (3,4).

Although the effect of oral anticoagulants on the frequency of stroke is clear, its impact on the severity of stroke and stroke-related inpatient outcomes among patients with AF has been less well studied, particularly in the DOAC era. Previous studies from the VKA era suggest that strokes happening while on anticoagulation may not have as severe short and long-term clinical outcomes (5–9). Few other studies in the DOAC era have shown similar outcomes (9–13). However, contemporary data on the inpatient outcomes of acute ischemic stroke among anticoagulated patients with AF in the United States in the DOAC era is scarce. Therefore, we set to use a nationwide, representative database to compare the inpatient outcomes of acute ischemic stroke among patients with AF on long-term AC compared to nonusers in the US.

## Methods

### Study Data

We utilized a retrospective study using the National Inpatient Sample (NIS) database. This database is the largest source of inpatient care data in the United States, compiled from billing information provided by hospitals to state data organizations as part of the Healthcare Cost and Utilization Project (HCUP). It includes over 100 clinical data elements from over 7 million unweighted hospital admissions annually, representing 20% of all U.S. hospital admissions. We applied a standardized sampling and weighting methodology, as provided by the HCUP, to make national estimates for the entire hospitalized population in the U.S. The NIS database is widely used for assessing inpatient outcomes in various conditions and procedures, including AF and ischemic stroke (14–16).

The study was exempted from institutional review board approval due to the use of a deidentified administrative database. We strictly followed the NIS survey methodology for data interpretation, research design, and analysis, in accordance with HCUP guidelines. The study also conformed to the Strengthening the Reporting of Observational Studies in Epidemiology Statement guidelines and incorporated best practice methodologies for using claims datasets.

### Study Population and Outcomes

We identified hospitalizations with primary diagnosis codes for acute ischemic stroke and secondary diagnosis codes for AF from 2016 to 2020, based on the *International Statistical Classification of Diseases and Related Health Problems, Tenth Revision (ICD-10)* codes. Inclusions were stratified by patients on long-term AC. All utilized ICD-10 diagnosis and procedure codes are listed in Supplementary Table 1.

For each hospitalization, we collected data on patient demographics, clinical characteristics, and hospital characteristics. Demographic variables included were age, sex, and race/ethnicity. Hospital characteristics included location/teaching status (rural, urban nonteaching, or urban teaching) and hospital bed size (small, medium, or large), while clinical characteristics covered relevant co-morbidities. The ICD-10-CM codes used for these variables have been previously validated and are consistent with prior publications (14–17).

The primary outcome of interest was all-cause in-hospital mortality. Secondary outcomes included stroke severity by National Institutes of Health Stroke Scale (NIHSS), intravenous thrombolysis and mechanical thrombectomy rates, in-hospital bleeding complications, hospital length of stay (LOS), total hospital costs, and discharge disposition. Patients with an NIHSS score of 1 to 4 were categorized as mild, 5-15 as moderate; 16 to 20 as moderate to severe, and above 21 as severe (18). The corresponding ICD-10 codes for each in-hospital outcome were identified using the same method as for co-morbidity codes.

### Statistical Analysis

We compared categorical variables using the Pearson Chi-Squared test and continuous variables using the Student t-test for normally distributed data, and the Mann-Whitney U test for non-normally distributed data. Categorical data were presented as counts and percentages, and continuous data were expressed as median with interquartile ranges (IQRs).

Multivariate analysis provided adjusted odds ratios (aORs) for binary variables, controlling for age, sex, and significant comorbidities. Associations were expressed through aORs with 95% confidence intervals (CI). The variables for adjustment in the multivariate regression model were chosen in advance based on their potential impact on in-hospital outcomes (see Supplementary Table 3). Statistical significance was set at p < 0.05. All analyses were performed using SPSS (IBM SPSS Statistics, Version 28.0). Weighted data were utilized for all statistical analyses, and missing demographic data were addressed through multiple imputation to the dominant category as recommended by the HCUP.

## Results

A total of 655,540 patients with AF hospitalized with a primary diagnosis of acute ischemic stroke were identified between 2016 and 2020. Of these, 194,560 (29.7%) were on long-term AC (**Figure 1)**. AC users tended to be slightly younger (mean age ± SD, 77± 10 vs. 78 ± 11 years; p <0.001), predominantly female (50.5% vs. 54.2%; p <0.001), and more frequently White (77.5% vs. 76.9%; p <0.001). They were also more likely to have persistent (4.5% vs 3.6%, p <0.001) and permanent AF (31.9% vs 17.8%, p <0.001) compared to AF patients not on AC. On the other hand, AC nonusers were more likely to have paroxysmal AF (36.2% vs 29.7%, p <0.001). Patients on AC had a higher mean CHA₂DS₂-VASc score (mean ± SD, 4.48±1.6 vs 4.20±1.6, p <0.001), and a higher proportion of patients with a CHA₂DS₂-VASc score between 5 and 9 (48.1% vs 41.2%, p<0.001). Patients on AC were less likely to be admitted to large (53.9% vs 55.2%, p <0.001), urban teaching hospitals (71.9% vs 74.0%, p <0.001) compared to patients not on AC. AF Patients on previous AC had a higher baseline comorbidity burden such as hyperlipidemia (64.3% vs 58.9%, p <0.001), hypertension (90.8% vs. 88.2%; p <0.001), diabetes mellitus (37.4%, vs. 35.1%; p <0.001), heart failure (33.8% vs. 29.9%; p < 0.001), chronic kidney disease (24.0% vs. 23.5%; p < 0.001), and coronary artery disease (36.2% vs. 32.1%; p <0.001), and were more likely to have had a previous stroke or TIA (35.1% vs 24.6%, p <0.001) compared to AF patients not on AC. The baseline characteristics are listed in **Table 1**.

**Figure 1.**
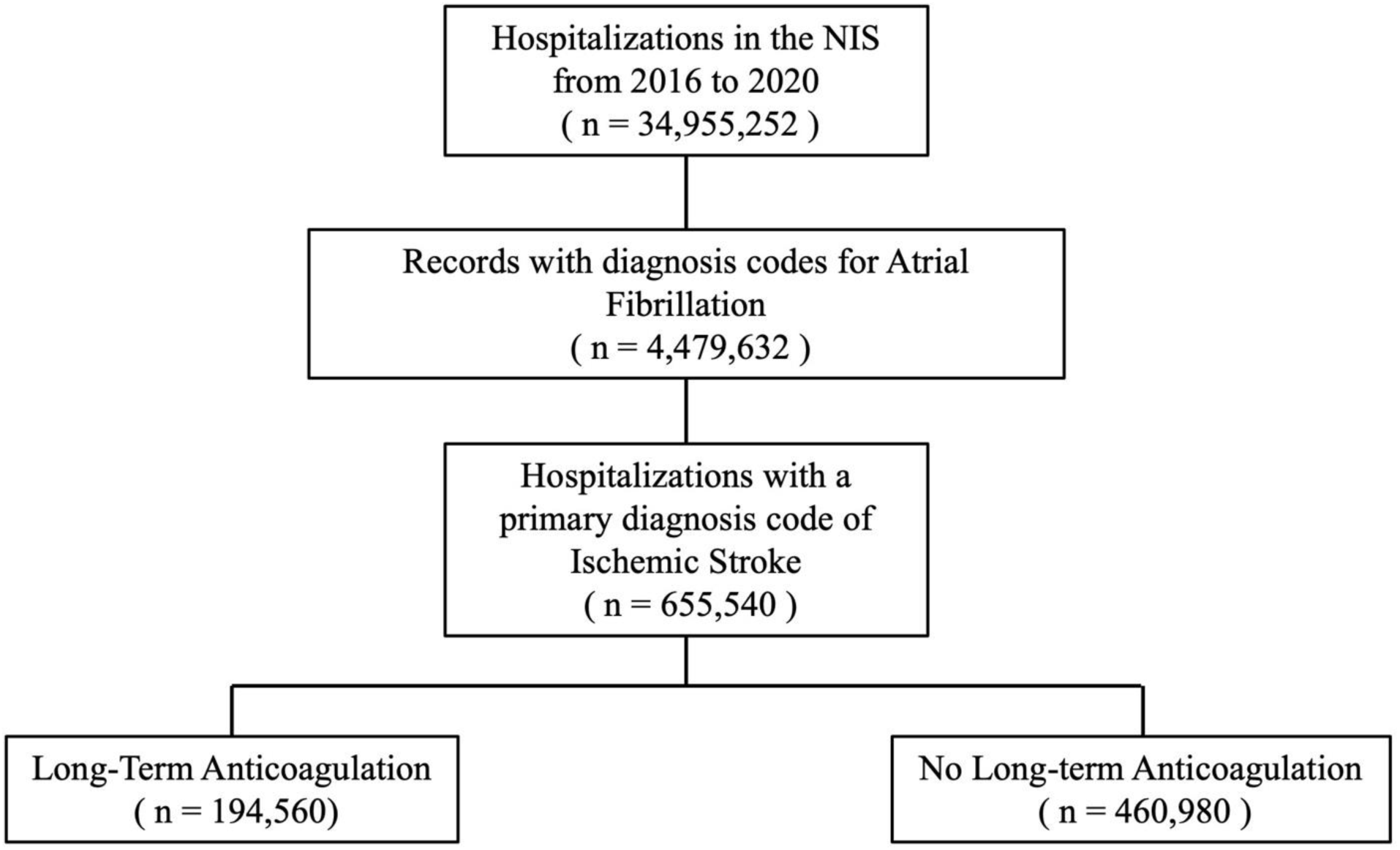
Study flow diagram showing inclusion and exclusion criteria. Hospitalization counts represent national-level estimates.

**Table 1.** Baseline characteristics in patients undergoing pericardiocentesis, with and without pulmonary hypertension.

|  | No Anticoagulation, n=460,980 (%) | Anticoagulation, n=194,560 (%) | P |
| --- | --- | --- | --- |
| Age, years [ IQR ] | 80 [ 71- 87 ] | 79 [ 71 - 86 ] | < 0.001 |
| Female | 249,720 (54.2) | 98,345 (50.5) | < 0.001 |
| Race |  |  |  |
| White | 343,670 (76.9) | 146,480 (77.5) | < 0.001 |
| Black | 46,745 (10.5) | 20,285 (10.7) |  |
| Hispanic | 30,030 (6.7) | 12,215 (6.5) |  |
| Other Races | 26,435 (6.0) | 10,052 (5.4) |  |
| **Atrial Fibrillation type** |  |  |  |
| Paroxysmal | 166,855 (36.2) | 57,735 (29.7) | <0.001 |
| Persistent | 16,630 (3.6) | 8,815 (4.5) |  |
| Permanent | 82,275 (17.8) | 62,035 (31.9) |  |
| Unspecified | 195,220 (42.3) | 65,975 (33.9) |  |
| Mean CHADSVASc<br>±SD | 4.20 ±1.57 | 4.48 ± 1.56 | <0.001 |
| CHADSVASc 0 - 1 | 20,920 (4.5) | 4,875 (2.5) |  |
| CHADSVASc 2 - 4 | 250,240 (54.3) | 96,005 (49.3) |  |
| CHADSVASc 5 - 9 | 189,820 (41.2) | 93,680 (48.1) |  |
| **Hospital Characteristics** |  |  |  |
| Relative bed size category of hospital |  |  |  |
| Small | 74,800 (16.2) | 33,810 (17.4) | < 0.001 |
| Medium | 131,905 (28.6) | 55,950 (28.8) |  |
| Large | 254,275 (55.2) | 104,800 (53.9) |  |
| Location/teaching status of hospital |  |  |  |
| Rural | 31,795 (6.9) | 15,360 (7.9) | < 0.001 |
| Urban nonteaching | 88,005 (19.1) | 39,375 (20.2) |  |
| Urban teaching | 341,180 (74.0) | 139,825 (71.9) |  |
| Hospital Region |  |  |  |
| Northeast | 87,700 (19.0) | 36,295 (18.7) | < 0.001 |
| Midwest | 98,130 (21.3) | 47,120 (24.2) |  |
| South | 181,465 (39.4) | 72,840 (37.4) |  |
| West | 93,685 (20.3) | 38,305 (19.7) |  |
| **Comorbidities** |  |  |  |
| Coagulopathy | 9,255 (2.0) | 5,845 (3.0) | <0.001 |
| Hypertension | 406,525 (88.2) | 176,625 (90.8) | <0.001 |
| Hyperlipidemia | 271,480 (58.9) | 125,115 (64.3) | <0.001 |
| Diabetes | 160,385 (35.1) | 72,140 (37.4) | <0.001 |
| Hypothyroidism | 86,105 (19.0) | 37,480 (19.5) | <0.001 |
| Obesity | 59,500 (12.9) | 26,750 (13.7) | <0.001 |
| Tobacco use | 140,805 (30.5) | 70,325 (36.1) | <0.001 |
| Alcohol use | 15,960 (3.5) | 4,915 (2.5) | <0.001 |
| Chronic lung disease | 83,575 (18.1) | 37,180 (19.1) | <0.001 |
| Chronic liver disease | 11,165 (2.4) | 3,760 (1.9) | <0.001 |
| CKD | 107,675 (23.5) | 46,585 (24.0) | <0.001 |
| Dialysis dependent | 6,665 (1.4) | 3,040 (1.6) | <0.001 |
| Anemia | 72,870 (15.8) | 27,475 (14.1) | <0.001 |
| CAD | 147,915 (32.1) | 70,430 (36.2) | <0.001 |
| Heart Failure | 137,830 (29.9) | 65,845 (33.8) | <0.001 |
| Peripheral vascular disease | 42,810 (9.3) | 17,830 (9.2) | 0.118 |
| Valve disease | 9,245 (2.0) | 11,855 (6.1) |  |
| Malignancy | 22,965 (5.0) | 10,095 (5.2) | <0.001 |
| **Previous History** |  |  |  |
| History of MI | 540,035 (8.7) | 19,545 (10.0) | 0.125 |
| History of TIA/Stroke | 113,135 (24.6) | 68,315 (35.1) | < 0.001 |
| History of Medication Noncompliance | 23,080 (5.0) | 15,735 (8.1) |  |

*IQR = interquartile range, SD = Standard deviation, CKD = Chronic Kidney Disease, CAD = Coronary Artery Disease, MI = Myocardial Infarction, TIA= Transient Ischemic Attack*

**Table 2.** Ischemic stroke characteristics and severity in atrial fibrillation patients, stratified by long-term AC use.

|  | **No**<br>**Anticoagulation**<br>**n= 460,980 (%)** | **Long-term**<br>**Anticoagulation**<br>**n= 194,560 (%)** | **P value** |
| --- | --- | --- | --- |
| **Stroke mechanism, n (%)** |  |  |  |
| Embolic | 155,130 (33.7) | 59,495 (30.6) | < 0.001 |
| Thrombotic | 30,735 (6.7) | 10,335 (5.3) | < 0.001 |
| Other/unspecified | 275,115 (59.6) | 124,730 (64.1) | < 0.001 |
| **Vessel occluded, n (%)** |  |  |  |
| Anterior Cerebral Artery | 9,975 (2.2) | 3,955 (2.0) | < 0.001 |
| Middle Cerebral Artery | 166,055 (36.0) | 58,380 (30.0) | < 0.001 |
| Posterior Cerebral Artery | 25,000 (5.4) | 9,450 (4.9) | < 0.001 |
| Vertebral and Basilar | 9,600 (2.1) | 3,430 (1.8) | < 0.001 |
| Carotids | 23,000 (5.0) | 9,500 (4.9) | 0.069 |
| Other/ unspecified | 219,340 (47.6) | 104,640 (53.7) | < 0.001 |
| Small vessel | 8,010 (1.7) | 5,205 (2.7) | < 0.001 |
| **Stroke severity** |  |  |  |
| Median NIHSS score | 7 [ 2 – 16 ] | 5 [ 2 – 12 ] | < 0.001 |
| Mild (NIHSS score 1–4) | 70,885 (15.4) | 37,955 (19.5) | < 0.001 |
| Moderate (NIHSS score 5–15) | 63,685 (13.8) | 27,485 (14.1) | < 0.001 |
| Moderate-severe (NIHSS score 16–20) | 19,425 (4.2) | 6,630 (3.4) | < 0.001 |
| Severe (NIHSS score 21–42) | 26,175 (5.7) | 8,760 (4.5) | < 0.001 |

*NIHSS = National Institutes of Health Stroke Scale, SNF = Skilled Nursing Facility, ICF = Intermediate Care Facility, LOS = Length of Stay*

While the majority of strokes were unclassified (**Table 3**), a lower percent of strokes in patients on AC was attributed to embolic source, compared to patients not on AC (30.6% vs 33.7%, p <0.001). Vessels occluded in AC users were less often large vessels such as the middle cerebral (30.0% vs 36.0%, p <0.001), anterior cerebral (2.0% vs 2.2%, p <0.001), or posterior cerebral arteries (4.9% vs 5.4%, p <0.001) compared to nonusers. Furthermore, patients on AC often had small vessel occlusion (2.7% vs 1.7%, p <0.001) compared to patients not on AC. Median NIHSS score among AC users was lower (5 [2 – 12] vs 7 [2 – 16], p <0.001). They also had a higher proportion of strokes categorized as mild (NIHSS score ≤ 4) compared to nonusers (19.5% vs 15.4%, p <0.001), and less frequently underwent revascularization with intravenous thrombolysis (7.3% vs 16.6%, p < 0.001) or mechanical thrombectomy (7.1% vs 8.9%, p < 0.001).

**Table 3.** In-hospital mortality and outcomes of atrial fibrillation patients admitted with acute ischemic stroke, stratified by long-term AC use.

|  | **No Anticoagulation**<br>**n= 460,980 (%)** | **Long-term**<br>**Anticoagulation**<br>**n= 194,560 (%)** | **Adjusted\***<br>**OR [95% CI]** | **P value** |
| --- | --- | --- | --- | --- |
| **Complications, n (%)** |  |  |  |  |
| In-hospital Mortality | 36,030 (7.8) | 9,405 (4.8) | 0.62 [ 0.60 – 0.63 ] | < 0.001 |
| Intravenous Thrombolysis | 76,640 (16.6) | 14,260 (7.3) | 0.42 [ 0.41 – 0.43 ] | < 0.001 |
| Mechanical Thrombectomy | 40,835 (8.9) | 13,790 (7.1) | 0.85 [ 0.83 – 0.87 ] | < 0.001 |
| **Bleeding complications n (%)** |  |  |  |  |
| ICH | 42,530 (9.2) | 12,455 (6.4) | 0.69 [ 0.67 – 0.71 ] | < 0.001 |
| GI bleed | 9,810 (2.1) | 2,895 (1.5) | 0.74 [ 0.70 – 0.77 ] | < 0.001 |
| All major bleed | 59,190 (12.8) | 18,530 (9.5) | 0.73 [ 0.71 – 0.74 ] | < 0.001 |
| **Discharge Disposition, n (%)** |  |  |  |  |
| Routine | 103,525 (22.5) | 50,150 (25.8) | 1.26 [ 1.24 – 1.28 ] | < 0.001 |
| SNF or ICF | 232,825 (50.5) | 93,575 (48.1) | 0.90 [ 0.89 – 0.91 ] | < 0.001 |
| **Resource utilization, median [ IQR ]** |  |  |  |  |
| Mean LOS, days | 4 [ 3 – 7 ] | 4 [ 2 – 6 ] |  | < 0.001 |
| Mean total charge, USD | \$48,130 [ \$27,089 - \$91,45 ] | \$40,135 [ \$24,130 - \$74,378 ] | | < 0.001 |
| *Matching for age, sex, elective admission status, presence of cardiac tamponade, coagulopathy, obesity, hypertension, hypothyroidism, diabetes, CKD, liver disease, lung disease, anemia, congestive heart failure, valvular disease, CAD, peripheral artery disease, atrial fibrillation, smoking, alcohol use, history of stroke/TIA/ MI, and medication non-compliance. |  |  |  |  |

*ICH = Intracranial Hemorrhage, GI = Gastrointestinal. SNF = Skilled Nursing Facility, ICF = Intermediate Care Facility, LOS = Length of Stay*

Unadjusted in-hospital mortality rates among AF patients on long-term AC were significantly lower compared to those not on AC (4.8% vs 7.8%, p < 0.001). Bleeding complications such as intracranial hemorrhage (ICH) (6.4% vs 9.2%, p < 0.001) and Gastrointestinal (GI) bleeding events (1.5% vs 2.1%, p < 0.001) were also significantly lower in AF patients on previous AC. Moreover, AC users had a shorter hospital LOS (median, [IQR] ; 4 [3 – 7]vs 4 [2 – 6]; p < 0.001), and lower average total costs (median USD [IQR]; $48,130 [$27,089 - $91,45] vs $40,135 [$24,130 - $74,378]; p <0.001) compared to nonusers. AF patient on previous AC were discharged at lower rates to a skilled nursing facility (SNF) or intermediate care facility (48.1% vs 50.5%; p <0.001) and discharged at greater rates to home (25.8% vs 22.5%; p <0.001) compared to those not on AC.

After adjusting for potential confounders using a multivariate logistic regression, long-term AC use was associated with decreased odds of in-hospital mortality (aOR [95% CI]: 0.62 [0.60 - 0.63], p <0.001), intravenous thrombolysis use (0.42 [0.41 - 0.43], p <0.001), mechanical thrombectomy use (0.85 [0.83 - 0.87], p <0.001), ICH (0.69 [0.67 - 0.70], p <0.001), GI bleed (0.74 [0.70 - 0.77], p <0.001), and all major bleeding events (0.73 [0.71 - 0.74], p <0.001). Adjusted odds of routine discharge were significantly higher for patients on long-term AC (1.26 [1.24 - 1.28], p <0.001), while the odds of discharge to SNF or other type of intermediate care facility were lower (0.90 [0.89 - 0.91], p <0.001). Forest plot showing odd ratios of the unadjusted and adjusted analyses is shown in **Figure 2**.

**Figure 2.**
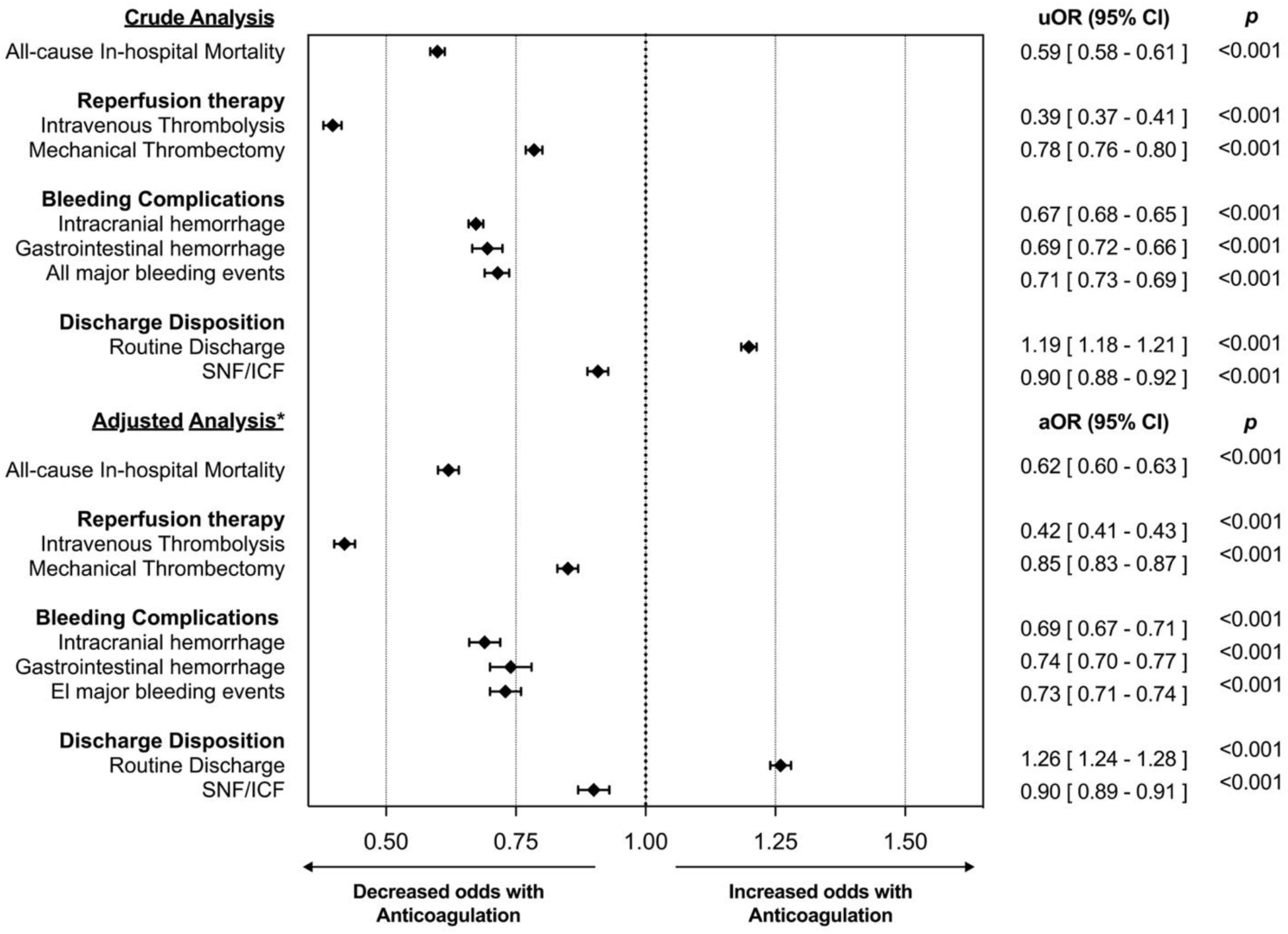
Forest plot showing crude and adjusted acute ischemic stroke outcomes in patients with atrial fibrillation on long-term anticoagulation versus patients not on long-term anticoagulation. *Adjusted analysis based on age, gender, race, and relevant co-morbidities. *uOR = unadjusted odds ratio; SNF = Skilled Nursing Facility; ICF = Intermediate Care Facility*.

## Discussion

In this study, we present several significant findings. Only 29.7% of AF patients who experienced an acute ischemic stroke were receiving long-term anticoagulation at the time of the event despite the availability of the newer direct acting oral anticoagulant drugs. Importantly, patients on anticoagulation had lower in-hospital mortality rate and reduced stroke severity. These strokes were less likely to involve major blood vessels and resulted in fewer discharges to skilled nursing facilities. Moreover, these patients had fewer chemical or mechanical thrombectomy procedures, and paradoxically, experienced fewer bleeding complications during their hospital stay.

Our data indicate that on third of AF patients were anticoagulated at the stroke onset, consistent with previous research (4,12,19,20). These patients typically had more comorbidities and higher average CHADS2VAS2C scores compared to those not on AC. Uncontrolled comorbidities, hyperlipidemia, high CHA2DS2VASc scores, and atherosclerosis have been identified as important risk factors for strokes despite AC therapy in AF patients (3,21). The more advanced AF disease process would likely have brought these patients to healthcare allowing more opportunity for providers to initiate guideline directed anticoagulation therapy. However, other factors such as increased AF burden (a sign of more advanced cardiac disease), insufficient anticoagulation (off label use of low dose DOAC or subtherapeutic VKA), or non-compliance are particularly influential in embolic breakthrough strokes (3,21,22). This aligns with our findings of a higher AF burden and medication non-compliance in those on previous AC.

The number of patients not on anticoagulants who might have previously undergone left atrial appendage occlusion (LAAO) in our sample is uncertain. However, as percutaneous LAAO was a relatively uncommon procedure during our study period, the relative number of patients who may have had prior LAAO, and thus benefit from embolic stroke prevention, is likely to be small. For patients where high bleeding risk is a concern and AC is contraindicated, LAAO could be beneficial. Among those not on AC, over 95% had a high-risk for stroke (CHADS2VAS2C ≥ 2) but were not reported to be on long-term AC. These findings indicate that a significant portion of strokes in AF patients may be attributable to insufficient or underutilized anticoagulation therapy. This highlights a potential avenue for reducing stroke rates among individuals with AF.

Using the large sample size from the NIS, this study demonstrated decreased stroke-associated in-hospital mortality and decreased stroke severity by NIHSS score in previously anticoagulated AF patients compared to those not on AC. Past multicenter studies have established that prior therapeutic warfarin treatment lessens stroke severity (5–9). A retrospective study by Hylek et al., initially revealed an association between successful anticoagulation and a decrease in disability as well as mortality within 30 days following an ischemic stroke (5). Comparable findings were observed in in-hospital data collected from a registry encompassing 11 hospitals in Canada (6). Studies also indicate that prior DOAC treatment lowers stroke severity and improves short-term outcomes, (9–13). A study spanning 2012 to 2015 demonstrated that therapeutic anticoagulation with warfarin or DOACs correlated with lower chances of moderate or severe stroke and reduced in-hospital mortality, which is in line with our findings (9). Another cohort reported that prior DOAC treatment was associated with lower risk of severe neurological deficit (NIHSS score ≥11) and death or disability (mRS score ≥3) at discharge in ischemic stroke patients with atrial fibrillation (11). Reported median NIHSS scores range from 4 to 8 for AF patients on therapeutic anticoagulation, versus 7 to 11 for those not on anticoagulation or on non-therapeutic doses, matching our results (9,12,13). These prior studies had smaller samples or focused mostly on functional short and long-term outcomes. To our knowledge, the present study has the largest sample size to examine contemporary, in-hospital outcomes in AF patients on previous AC admitted for acute ischemic stroke in the DOAC era.

The diminished stroke severity observed in patients on anticoagulation likely reflects smaller thrombus size, and thus decreased LVO and infarct volume (7,13,23–25). Decreased frequency of embolic strokes, which characteristically have a higher affinity for the total anterior circulation and often result in more severe strokes, is also thought to play a role (23–25). The literature is inconsistent regarding the predominant stroke etiology between embolic or non-embolic in AF patients that suffer strokes while on anticoagulation. In this study, embolic etiology was a primary mechanism for stroke. (12,22,26). Nevertheless, we observed lower involvement of total anterior circulation vessels (anterior and middle cerebral arteries), more small vessel circulation strokes, and fewer total embolisms in the anticoagulated group compared to those not on anticoagulation, indicating effective anticoagulation might lead to smaller embolus size and thus occlusion of smaller arteries (7,13,23–25).

In comparison to VKA, DOACs have a shorter half-life (6–14 hours) and a rapid onset of action (27). Current major stroke guidelines advise against routine intravenous thrombolysis within 48 hours of DOAC intake unless specific DOAC coagulation assays are normal (28,29). Many patients on DOACs, despite being eligible, do not receive intravenous thrombolysis due to the risk of hemorrhagic transformation (3,4,25). Similarly, we noted lower rates of intravenous thrombolysis in our observations. Mechanical thrombectomy rates were also lower among patients on AC. This differs from prior studies reporting increased rates of mechanical thrombectomy in this population as a reperfusion strategy since of intravenous thrombolysis is usually avoided (3,4,20,25). As guidelines recommend mechanical thrombectomy particularly in the setting of acute ischemic stroke and concomitant LVO (29), perhaps as anticoagulated patients presented with LVO less frequently, they required mechanical thrombectomy less often.

## Limitations

There are several limitations to this study. Data collected from the NIS database is primarily compiled based on ICD codes, which represent “claims data” for reimbursement purposes and for summarizing clinical presentations in retrospect. Thus, patient level data is not available, such as time from stroke onset to initial medical care, contraindications to IS treatments, or neuroimaging data. There could be unmeasured and unknown confounding factors that are not captured in the NIS database, such as patient preference and provider-level rationale, that are important determinants of clinical decisions and may in part account for observed associations. We did not have data as to how many patients presented with newly diagnosed AF which is another confounder. As with all administrative datasets, misclassification bias is a limitation, although our use of previously validated ICD-10-CM codes for stroke in the primary discharge diagnosis position limits that bias for the identification of patients. It is more likely that we under captured patients under long-term AC, because of lack of documentation. When looking at stroke severity, 60.2% had missing NIHSS score. Since every observation represents a hospitalization, information before or after the hospital admission is not available, such as timing from stroke to admission, or 30-day outcomes. We also lack granularity in many of our variables due to the administrative nature of NIS. For example, information on the specific anticoagulant patients were taking or INR levels on admission (if on warfarin), and further ischemic stroke type classification are not available.

## Conclusion

In summary, using a large, nationwide database, this study supports a compelling argument that anticoagulation reduces stroke severity and stroke associated disability for those who have strokes despite anticoagulation. In particular, we observed that AC use in AF patients prior to stroke was associated with lower in-hospital mortality, lower stroke severity, and lower bleeding complications compared to patients not on AC. These patients had a shorter LOS and were less likely to be discharged to a SNF or ICF.

## Data Availability

The National Inpatient Sample is publicly available for purchase through the Agency of Healthcare Research and Quality. The ICD10 codes used in this study are available in the Supplementary Materials.

https://hcup-us.ahrq.gov/nisoverview.jsp

## Abbreviations

AF: Atrial Fibrillation
NVAF: Nonvalvular Atrial Fibrillation
AC: Anticoagulation
DOAC: direct oral anticoagulants
VKA: vitamin K antagonists
LAAC: Left Atrial Appendage Closure
IS: Ischemic Stroke
SEE: Systemic Embolic Events
TIA: Transient Ischemic Attack
NIS: National Inpatient Sample

